# Improving air quality and health in Northern Italy: limits and perspectives

**DOI:** 10.1101/2021.12.31.21268581

**Authors:** Angelo Robotto, Secondo Barbero, Roberto Cremonini, Enrico Brizio

## Abstract

A better air quality has led to a significant reduction of premature deaths over the past decade in Europe, as emissions of many pollutants declined considerably in the EU-27 Member States: SOx emissions by 76%, NOx by 42%, NMVOCs by 29% and PM2.5 by 29%. The present paper reports an in-depth analysis of the reasons why the regions of the Po valley, Northern Italy, still have difficulties to comply with EU air quality standards, in particular for PM10 and NO2, in spite of strong emission reductions carried out through careful Air Quality Plans put in practice during the last 2 decades. The analysis includes a consistent comparison of emission inventories for different European regions in Italy, Germany and Poland, the measured air quality trends and PM source apportionment in these areas, and, most of all, a thorough investigation of meteorological parameters influencing atmospheric pollutant dispersion and transport. The study reports that in the colder seasons, wind speed, PBL height and atmospheric pressure occurring in the Po basin are three to five times less efficient in diluting and dispersing pollutant if compared to regions north of the Alps. Due to the extremely disadvantageous orographic and climatic configuration of the Po Valley, only radical emission reductions could bring air quality into EU limit values with a questionable cost-benefit ratio of due policies. Provided that air quality standards (particularly for PM10 and PM2.5) aim at protecting people from adverse health effects arising from air pollution, it is however necessary to also consider the toxicity of atmospheric particulate in addition to PM10/PM2.5 mass concentration as a limit value. Based on existing toxicological studies and reports, a discussion is reported about PM toxicity factor depending on toxicity scores for source-specific aerosols and PM composition determined by Source Apportionment. Provided that PM components’ profiles are strongly different across Europe, the obtained PM toxicity factors range from 0.3 (for areas where the main PM contribution is referable to sea salts or inorganic matter) to 3.5 (where Elemental and Organic Carbon prevail), suggesting that, even at the same mass concentration, the effects of PM10/2.5 on human health are significantly variable and limit values should take into account differential toxicity.

Modern PM Source Apportionment techniques, along with reliable toxicity and epidemiological analyses, represent the right tools to overcome the shortcomings of the current regulation standard and build a new consistent health metric for ambient PM in the future, helping policy makers impose effective air quality measures to protect people health.

## 1. Introduction

The report ‘Air quality in Europe’ for 2020 by the European Environmental Agency (EEA, 2020a) shows that better air quality has led to a significant reduction of premature deaths over the past decade in Europe. Since 2000, emissions of atmospheric pollutants from transport strongly reduced, in particular for nitrogen oxides (NOx), even though mobility has grown up significantly in the same period.

Pollutant emissions from energy production showed large reductions as well, while emissions from buildings and agriculture could be still improved. EEA estimates that <<around 60,000 fewer people died prematurely due to fine particulate matter pollution in 2018, compared with 2009. For nitrogen dioxide, the reduction is even greater as premature deaths have declined by about 54 % over the last decade>>.

As a matter of fact, anthropogenic emissions of the main air pollutants that are NH_3_, non-methane volatile organic compounds (NMVOCs), nitrogen oxides (NOx), particulate matter (PM10 and PM2.5) and sulphur oxides (SOx), contribute to air quality problems in Europe, causing impacts on human health, vegetation and ecosystems.

Between 2005 and 2019, according to the commitments of the National Emission Ceilings Directive, emissions of many pollutants declined considerably in the EU-27 Member States: SOx emissions by 76%, NOx by 42%, NMVOCs by 29% and PM2.5 by 29%. The reductions have been strongly addressed by sector-specific EU legislation, such as the Industrial Emissions Directive, the Large Combustion Plants Directive and Euro standards for vehicles. NH_3_ emissions also reduced, but by only 8% overall, meaning that agriculture should still improve its environmental performance (EEA, 2021a).

Table 1 reports emission reductions for the main air pollutants in the 7 most populous country in EU obtained from 2005 to 2019, as pointed out by EEA. On average, Italy shows the second-best emissive improvement (−39%) after France (−44%), whereas Poland and Germany reached much lower reductions (respectively -26% and -27%).

**Table 1:** Emission reduction of the main air pollutants by Member State from 2005 to 2019

|  | NH3 | NMVOC | NOx | PM2.5 | SOx | average reduction 2005-2019 |
| --- | --- | --- | --- | --- | --- | --- |
| Germany | -3% | -25% | -31% | -33% | -45% | -27% |
| France | -5% | -40% | -48% | -51% | -78% | -44% |
| Italy | -15% | -33% | -51% | -20% | -74% | -39% |
| Spain | -2% | -23% | -52% | -8% | -88% | -35% |
| Poland | -6% | -16% | -23% | -21% | -62% | -26% |
| Romania | -17% | -29% | -34% | -7% | -84% | -34% |
| Netherlands | -19% | -11% | -43% | -46% | -66% | -37% |

Despite the efforts made to reduce emissions in EU, Bulgaria, Croatia, Czechia, Italy, Poland, and Romania still exceeded the European Union’s limit value coming from EU Air Quality Directives 2008/50/EC and 2004/107/EC for fine particulate matter (PM2.5) in 2018 and only four countries in Europe — Estonia, Finland, Iceland and Ireland — had fine particulate matter concentrations below the World Health Organization’s (WHO) stricter guideline values.

In Italy, in 2018, the percentage of urban population exposed to concentration above PM10 EU daily limit value (50 µg/m^3^ not to exceeded for more than 35 days per year, that is 90.41 percentile of daily average in a calendar year) in 2018 is still around 34.4%, 1.5% for PM2.5 annual limit value (25 μg/m^3^), 7.3 % for NO2 annual limit value (40 μg/m^3^).

In particular, the Po Valley, located in Northern Italy at the foot of the Alps, is characterized by a high density of anthropogenic emissions and the frequent occurrence of stagnant weather conditions. The area is known to be a “hot spot” for air quality where pollutant levels are still challenging despite a constant reductions of air pollutants’ emission in the last decade (Thunis et al., 2009). The Po basin is made up of 4 big Italian regions, Piedmont, Lombardy, Emilia Romagna and Veneto, with around 23.5 million inhabitants; the main features of this area are a strong industrialization and intensive farming and agriculture, producing 48% of Gross Domestic Product (GDP) of Italy in 2017. The 4 regions together emit 327 kt/y of NOx, 237 kt/y of ammonia, 60 kt/y of PM10 and 50 kt/y of PM2.5, 36 kt/y of SO_2_.

Because of the morphological characteristics of the Po Valley, that is closed on three sides by Alps and Apennines, pollutants’ background concentrations remain high for long periods during the cold season, with a large part of the particulate matter being due to secondary production. The Po basin represents the largest European area characterized by such geographical and meteorological adverse conditions that caused the partial failure of air quality remediation policies, the so-called regional Air Quality Plans, in the last two decades. Indeed, the concentrations of PM10 strongly decreased in the last twenty years but, although reducing to a quarter of what they measured in 2000, in some agglomerations and areas they still do not comply with the air quality limits.

In 2013, the regional governments of the area signed the first Po Valley Agreement, aiming at developing and coordinating short and long-term measures to improve the air quality of the Po Valley, focusing their actions on biomass domestic burning, transportation of goods and passengers, agriculture. In 2017, a second Agreement was approved, reinforcing the actions to reduce emissions.

On 10^th^ November 2020 (case C-644/18), the Court of Justice of the European Union declared that the Italian Republic failed to fulfil its obligations under the provisions of Article 13 of, in conjunction with Annex XI to, Directive 2008/50/EC of the European Parliament and of the Council of 21^st^ May 2008 on ambient air quality and cleaner air for Europe by having systematically and continuously exceeded, from 2008 to 2017, the daily and annual limit values applicable to PM10 concentrations in specific areas, highlighting that the exceeding is “still in progress”. With the same sentence, the Court of Justice found that the Italian Republic has also failed to comply with the obligation established by art. 23, in conjunction with Annex XV of Directive 2008/50/EC, by failing to adopt as from 11^th^ June 2010 appropriate measures to ensure compliance with the limit values for PM10 in all those zones and to ensure that the exceedance of the limit values is kept as short as possible.

Meanwhile, in December 2021, the European Commission started the revision of ambient air quality directives, which will also include a revision of the air quality standards to better align with the new guidelines of the World Health Organization, published in September 2021 (WHO, 2021).

According to the last news by the European Environment Agency (EEA, 2021b), the vast majority of EU citizens are exposed to levels of pollutants that can cause damage to health: 97% of the urban population is exposed to levels of fine particulate matter higher than those indicated in the new WHO guidelines, 94% for nitrogen dioxide and 99% for ozone.

Within the described framework, it is advisable to investigate the real effectiveness of the measures to be taken to respect air quality limits in Northern Italy, focusing on the health effects of the main air pollutants. In the next chapters, in-depth analyses about regional emission inventory across EU, PM source apportionment, role of secondary particulates, air quality trends observed during Covid-19 lockdown in Italy, influence of three key meteorological parameters on air quality will be reported. Then, a discussion about PM toxicity data and regulation standards is proposed as well.

## 2. Materials and methods

### 2.1 Emission inventories and air quality trends in EU

The first approach to address air quality problems consists in the analysis of emission inventories referring to different areas. The European Union emission inventory report 1990-2018 (EEA, 2020b) highlights EU emission trends over 28 years for the main pollutants, nitrogen oxides (NOx), non-methane volatile organic compounds (NMVOCs), sulphur oxides (SOx), ammonia (NH3), carbon monoxide (CO), particulate matter (PM10 and PM2.5), Heavy Metals, Persistent Organic Pollutants (POPs) and Polycyclic Aromatic Hydrocarbons (PAH). In the described document, national emission contributions of EU Member States are identified as well.

However, to develop coherent comparisons between different areas, emission inventories at the regional level are also required. In order to collect these data, regional emission inventories for Italian Northern regions, for North Rhine-Westphalia (NRW) in North Western Germany (the most populous State in Germany), Baden-Württemberg and Bavaria in Southern Germany, Lesser Poland and Silesian in Southern Poland, have been downloaded from the official regional Administrations sites (Regione Piemonte; Regione Lombardia; Regione Veneto; Regione Emilia Romagna; Landesamt für Natur, Umwelt und Verbraucherschutz Nordrhein-Westfalen; Landesanstalt für Umwelt, Messungen und Naturschutz Baden-Württemberg LUBW; Institut für Energiewirtschaft und Rationelle Energieanwendung; Lochno et al., 2020; Załupka et al, 2020). All inventories are recent and have been elaborated according to EEA European guidelines (EEA, 2019).

When evaluating air quality, it is important to understand the difference between primary pollutants, such as NOx, SOx or PM, and secondary pollutants, that are secondary PM (organic and inorganic secondary aerosols) and O_3_. De Leeuw (2002) developed a set of aerosol formation factors, that are fractions of gaseous primary pollutants converted to particulate matter: the author set the factors equal to 0.88 for NOx, 0.54 for SOx, 0.64 for NH3, whereas the contribution of VOC to the total PM10 emissions in EU was estimated to be less than 1.5%. Therefore, we focused our attention primarily on emission inventories for NOx, NH3, SO2 (gaseous precursor of PM), PM10, PM2.5 (where available) and CO2 equivalent emissions as an overall indicator of anthropization.

Table 2 reports emission inventory data together with population and surface in km^2^ of the studied Europeans regions. Regions to compare have been chosen according to equivalent or comparable surfaces, even though population density can be quite different. We developed the following emissive comparisons:

**Table 2:** Emissions of main atmospheric pollutants in the Po Basin (Piedmont+Lombardy+Emilia Romagna+Veneto) and other European regions

| EU State | Region/State | population | surface (km<sup>2</sup>) | CO<sub>2</sub>eq (kt/y) | NH<sub>3</sub> (t/y) | NO<sub>x</sub> (t/y) | PM<sub>10</sub> (t/y) | PM<sub>2.5</sub> (t/y) | SO<sub>2</sub> (t/y) | ref. year |
| --- | --- | --- | --- | --- | --- | --- | --- | --- | --- | --- |
| IT (NW) | Piedmont | 4.273.210 | 25.402 | 40.547,9 | 40.007,6 | 72.945,8 | 16.911,9 | 12.680,1 | 8.374,8 | 2015 |
| IT (NW) | Lombardy | 9.964.993 | 23.844 | 76.970,1 | 97.114,0 | 111.474,6 | 17.822,9 | 15.040,1 | 11.179,8 | 2017 |
| IT (NE) | Emilia Romagna | 4.448.545 | 22.446 | 34.605,4 | 48.438,1 | 81.173,2 | 10.958,4 | 9.532,4 | 11.435,3 | 2017 |
| IT (NE) | Veneto | 4.841.175 | 18.345 | 33.794,0 | 51.854,6 | 61.450,7 | 13.904,5 | 12.703,9 | 5.003,3 | 2017 |
| DE (NW) | North Rhine-Westphalia | 17.925.570 | 34.084 | 276.099,3 | 91.402,9 | 265.536,9 | 18.952,3 | / | 83.277,7 | 2016 |
| DE (SE) | Bavaria | 13.124.737 | 70.550 | 110.053,7 | 86.048,0 | 168.763,0 | 16.172,0 | / | 25.740,0 | 2010 |
| DE (SW) | Baden-Württemberg | 11.100.394 | 35.751 | 83.266,9 | 54.404,8 | 112.147,3 | 13.331,0 | / | 12.801,6 | 2018 |
| PL (S) | Lesser Poland | 3.404.863 | 15.183 | 25.105,7 | / | 53.247,3 | 32.633,2 | 28.231,9 | 42.694,6 | 2018 |
| PL (S) | Silesian | 4.524.091 | 12.333 | 59.730,8 | / | 83.842,3 | 36.283,5 | 31.070,2 | 36.897,7 | 2018 |

1. North-Eastern Italy (regions Veneto and Emilia Romagna, equivalent to 9,289,720 inhabitants and 40,791 km^2^ all) vs North Rhine-Westphalia (17,925,570 inhabitants and 34,084 km^2^);
2. Lombardy (9,964,993 inhabitants and 23,844 km^2^) vs Baden-Württemberg (11,100,394 inhabitants and 35,751 km^2^);
3. Po Valley (regions Piedmont, Lombardy, Veneto and Emilia Romagna, equivalent to 23,527,923 inhabitants and 90,037 km^2^) vs Southern Germany (regions Bavaria and Baden-Württemberg, that is 24,225,131 inhabitants and 106,302 km^2^);
4. Piedmont (4,273,210 inhabitants and 25,402 km^2^) vs Southern Poland (regions Lesser Poland and Silesian, equivalent to 7,928,954 inhabitants and 27,516 km^2^).

At the same time, official air quality data have been elaborated for the same areas where emission inventories were at disposal. We focused our attention on PM10 concentrations (90.41 percentile of daily concentration) and NO2 concentrations (yearly average), that are two critical pollutants for air quality. Five years of air quality statistics from 2015 to 2019 have been downloaded from EEA official website (https://www.eea.europa.eu/data-and-maps/dashboards/air-quality-statistics-expert-viewer); 2020 has been avoided because of the influence of Covid-19 lockdowns on air quality. PM10 and NO2 concentrations have been averaged over all the air quality stations operating within the reference area (rural, urban and suburban sites).

From the same official database, air quality trends over the period 2001-2019 have been obtained for three urban-traffic and three urban-background stations of the cities of Torino (886,837 inhabitants in Piedmont, IT), Düsseldorf (619,294 inhabitants in North Rhine-Westphalia, DE) and Krakow (766,683 inhabitants in Lesser Poland, PL).

### 2.2 Parameters influencing pollutant atmospheric dispersion and transport

Many meteorological parameters influence dilution, dispersion and transport of pollutants emitted into the atmosphere, including wind speed, wind direction, atmospheric pressure, and the atmospheric stability. Atmospheric stability is in turn described by different meteorological parameters, such as the height of the planetary boundary layer (PBLH) or mixing height, stability class, vertical temperature gradient, solar radiation, etc. (Stull, 1988). It is well known that when pollutant transformation mechanisms are not considered, the wind speed-pollutant concentration relationship is inverse. The depth of PBL is a key parameter in cumulus convection and it is crucial in determining near-surface atmospheric pollutant concentrations (Jacob, 1999). Indeed, PBLH determines the volume where emitted pollutants are diluted and dispersed and therefore it directly affects pollutants atmospheric concentrations. Daytime mixed-layer (ML) height is regarded as the location of a capping temperature inversion over the convective boundary layer. Several studies demonstrated a clear relationship between carbon dioxide and aerosol concentrations and ML depth through entrainment processes (Raupach et al., 1992; Denmead et al., 1996; Quan et al., 2013; Pal et al., 2014).

The depth of PBL is influenced by surface topography, incoming solar radiation, temperature, and local winds. The PBLH can be 1-2 km deep, and it impacts not only air quality but also climate and weather. It changes throughout the day as the ground heats up and cools down; it is also a strongly seasonal parameter (during the winter PBLH is lower). A higher PBL is generally better because emitted pollutants are more diluted; however, a higher PBL allows transportation of pollutants from one place to another.

Critical conditions for air quality are in most cases related to the expansion of a high-pressure pattern; this way, besides wind speed and PBL height, mean sea level pressure is a parameter of interest when studying air quality (Czernecki et al., 2016).

To investigate weather parameters affecting air pollutants’ concentrations, long-term re-analysis data have been accessed. ERA5 is the fifth generation ECMWF atmospheric reanalysis of the global climate: combining observations with model data, re-analysis constitutes a globally complete and consistent dataset (Hersbach et al., 2020). Spanning from 1979 to 2020, data are re-gridded to a regular latitude-longitude grid with 0.25° by 0.25° resolution. A complete overview of ERA5 suite is available at https://confluence.ecmwf.int/display/CKB/The+family+of+ERA5+datasets.

Here, monthly average diurnal pattern of PBLH, atmospheric mean sea level pressure and 10-meters wind speed relating to Essen (7.0E, 51.5N - North Rhine-Westphalia, Germany), Torino (7.7E, 45.0N - Piedmont, Italy) and Katowice (19.0E, 50.3N - Silesian, Poland), have been obtained from Copernicus Climate Data Store (https://cds.climate.copernicus.eu/#!/home). The meteorological and dispersive characteristics of the three localities and their regions are compared focusing on the coldest months of the year, November-February, over the whole period (1979-2020).

### 2.3 PM Source Apportionment in Northern Italy

The PM Source Apportionment (SA) represents the data collection and processing method that quantitatively determines the importance of individual contributions to ambient air pollutant concentration, in our case PM10 (JRC, 2014). Among the different approaches that can be used, the “receptor” analytical approach allows to obtain estimates starting from the chemical composition of PM10 sampled at significant sites from the point of view of the objective, applying specific statistic techniques such as the Positive Matrix Factorization (PMF) by U.S.EPA.

The choice of the analytes to be quantified in PM10 is made taking into account the need to define the main constituents of particulate as well as the tracer compounds of particular emissive sources (Levoglucosan for Biomass burning and Copper for traffic, for example). Cations (Na^+^, NH^4+^, K^+^, Mg^2+^) and anions (Cl^-^, NO2^-^, Br^-^, NO3^-^, PO4^3-^, SO4^2-^), elements (Al, Si, P, S, Cl, K, Ca, Ti, V, Cr, Mn, Fe, Ni, Cu, Zn, Br, Rb, Pb), the carbonaceous fraction (OC and EC, Organic and Elemental Carbon), polycyclic aromatic hydrocarbons (PAHs), are usually determined for PM Source Apportionment.

For SA, the determination of ions (cations and anions) is crucial to estimate the amount of secondary inorganic aerosols (SIA): nitrates and sulphates come from combustion (industry, traffic, and heating), ammonium derives mainly from agriculture and animal intensive rearing.

The Elemental Carbon is a PM fraction containing only C, not bound to other elements; it is a primary pollutant emitted during the incomplete combustion of fossil fuels and biomass and can be emitted from natural and anthropogenic sources in the form of soot. In urban areas it can be used as a tracer of internal combustion engines emissions and the wide range of chemical species associated, including organic compounds such as PAHs.

The Organic Carbon (OC) includes many compounds with different volatilities; it is both primary and secondary pollutant. The main sources of primary OC are the natural or anthropogenic combustion of biomass, fossil fuels (industry, transport, etc.) and biological material. The secondary OC can be formed by photochemical oxidation of volatile precursors (VOC). OC includes a large set of compounds where tetravalent carbon is chemically bound to other atoms, hydrogen, oxygen, sulphur, nitrogen, phosphorus, chlorine, etc.

Lastly, the crustal oxide compounds (Al, Si, Ca, etc.) present in PM10 are important to evaluate the resuspension contribution to PM pollution.

In the Po Valley, PM Source Apportionment has been carried out since 2013, in some specific site such as the urban-background stations of Milano-Pascal or Bologna-Gobetti (Perrino et al., 2020; EU LIFE IP Prepair, 2021).

The Source Apportionment has been also carried out for the stations of Torino-Lingotto (urban-background) and Revello-Staffarda (rural) in Piedmont (N-W Italy), referring to the period December 2016 - June 2017 (Regione Piemonte, 2018). In this case, the chemical analysis of the samples was followed by statistical pre-elaborations consisting in characterization of the soils in the monitoring sites, calculation of enrichment factors, study of the correlation between chemical parameters, exploration with cluster analysis, attribution of uncertainty; after that, the EPA PMF 5.0 (Positive Matrix Factorization) statistical model was applied based on the so-called fingerprint of the sources and their space-time variability, through the application of multivariate analysis techniques.

The analytical Source Apportionment developed for Piedmont (N-W Italy) represents a parallel technique strengthening the evaluations obtained with Source Apportionment modelling methods, starting from emission inventories and from the meteorological variables measured to simulate the chemical reactions that take place in the atmosphere.

### 2.4 Air quality during Covid-19 lockdown

In 2020 the regional Environmental Protection Agency for Piedmont (Arpa Piemonte) published a study (Arpa Piemonte 2020) focused on the link between the reduced atmospheric emissions due to the limiting measures following the COVID-19 emergency (mainly involving traffic and productive activities) and the ambient air pollutants’ concentrations (PM10 and nitrogen dioxide) in N-W Italy. The purpose was to verify the presence of an additional effect on the ordinary decrease of atmospheric pollutants’ concentration normally occurring with the start of the spring season. In this regard, it is important to underline that, unlike PM10, the concentrations of nitrogen oxides (NOx), mainly emitted by vehicular traffic, respond more directly to variations in emissions.

PM10 and NO2 emissions have been estimated for the period 01 January 2020-30 April 2020 (in Italy a hard lockdown started on March 9 2020); PM10 and NO2 concentrations for the first four months of 2020 were analysed by comparing them with those measured in the same period by the stations of the regional air quality network for the years 2012-2019.

## 3. Results

The emission inventories of different regions in Europe, as described in Section 2.2, are reported in Figure 1, 2, 3 and 4. Figure 1 demonstrates that, although North-Eastern Italy has lower (or at least comparable) atmospheric emissions, from a larger surface in addition, both for particulate matter and its gaseous precursors, the measured concentrations of PM10 are much higher than in North Rhine-Westphalia (NRW, North-Western Germany). In particular, 90.41 percentile of PM10 daily average in a calendar year is higher than EU air quality standards (50 μg/m^3^ not to exceeded for more than 35 days per year) in 4 out of 5 analysed years. On the contrary, NO2 yearly concentrations are higher for NRW than for Northern Italy, due to higher emissions and different secondary aerosol formation mechanisms.

**Figure 1:**
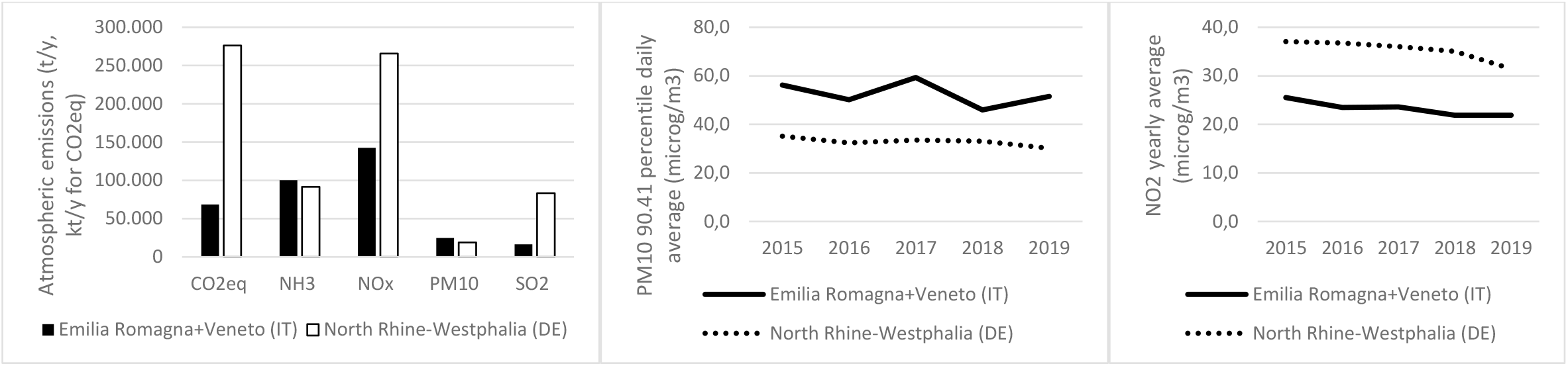
Emission inventory for North Eastern Italy (Emilia Romagna+ Veneto regions) and North Rhine-Westphalia in Germany; yearly NO2 concentrations and PM10 90.41 percentile daily average are reported for the period 2015-2019

**Figure 2:**
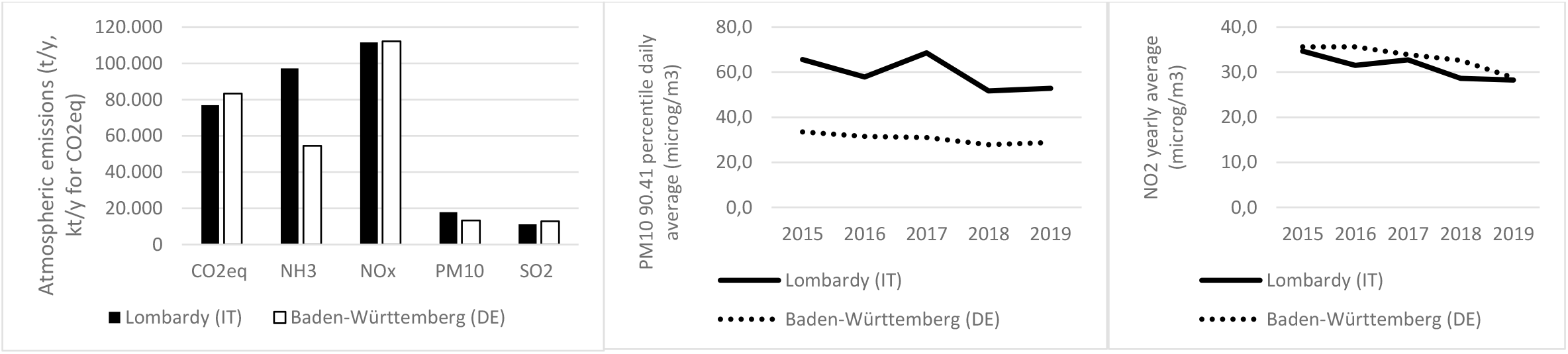
Emission inventory for Lombardy in Italy and Baden-Württemberg in Germany; yearly NO2 concentrations and PM10 90.41 percentile daily average are reported for the period 2015-2019

**Figure 3:**
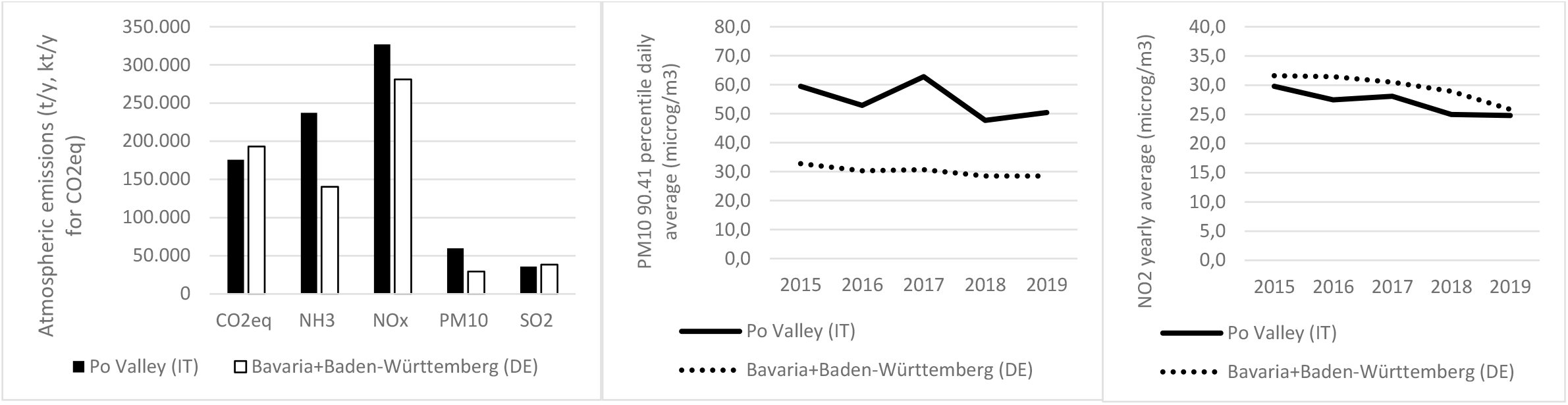
Emission inventory for Po Valley (Piedmont+Lombardy+Emilia Romagna+Veneto) in Italy and Southern Germany (Bavaria+Baden-Württemberg); yearly NO2 concentrations and PM10 90.41 percentile daily average are reported for the period 2015-2019

**Figure 4:**
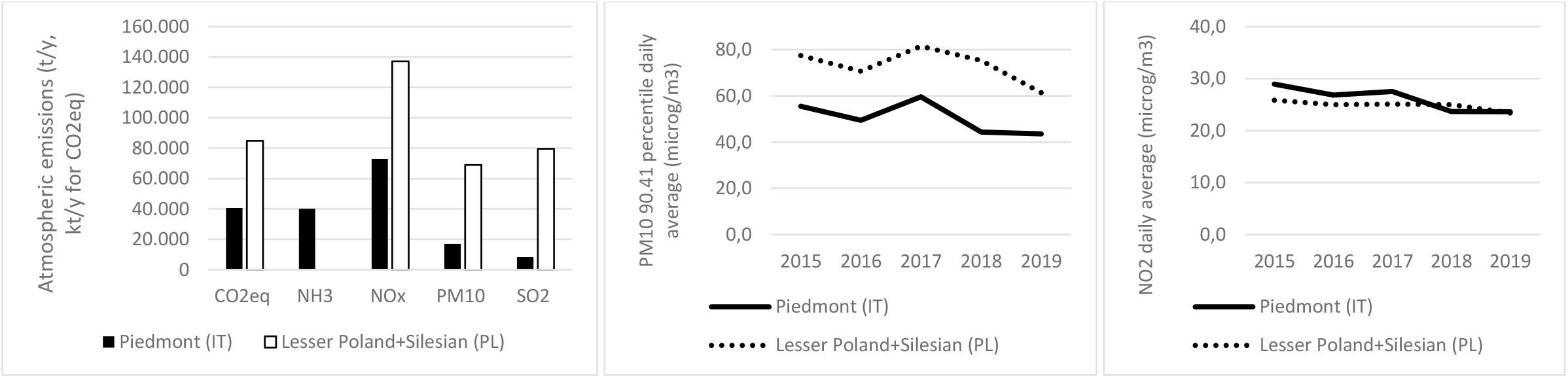
Emission inventory for Piedmont in Italy and Southern Poland (Lesser Poland and Silesian); yearly NO2 concentrations and PM10 90.41 percentile daily average are reported for the period 2015-2019

Figure 2 reports similar results for Lombardy (Northern Italy) and Baden-Württemberg (South-Western Germany): emissions are comparable (except for NH3 that is higher in Lombardy), but PM10 concentrations are much higher in Lombardy and NO2 almost equivalent. Figure 3, dealing with the entire Po Valley compared to Southern Germany, confirms the behaviours highlighted by Figure 2.

Finally, Figure 4 shows an interesting comparison between Piedmont (N-W Italy) and 2 regions of Southern Poland, with similar emissive surfaces. In this case, lower Poland has important emissions, relating to PM10 (4 times higher than for Piedmont), SO2 (10 times higher), NOx (twice those from Piedmont). Despite the stronger emissions, PM10 concentrations are just 45% higher in Poland, and NO2 turned out to be even lower than in Piedmont.

Figure 5 and 6 report air quality trends for PM10 concentrations (90.41 percentile daily average) in Torino (IT), Düsseldorf (DE) and Krakow (PL). The data highlight a decreasing trend for all considered sites, of different entities: concerning the urban-traffic stations, PM10 concentrations decreased by 52% in Torino, 49% in Krakow and 43% in Düsseldorf during the period 2001-2019; only Düsseldorf stably complies with PM10 daily limit value of 50 μg/m^3^. As far as urban-background stations are concerned, concentrations decreased by 52% in Torino and 36% in Düsseldorf in the period 2006-2019. It is interesting to observe that urban-traffic and urban-background PM10 concentrations in Torino show almost the same values, whereas in the other two studied cities urban-traffic stations report significantly higher concentrations with respect to background areas. Almost constant pollutant concentrations are generally due to stagnation conditions and poor atmospheric dispersion potential.

**Figure 5:**
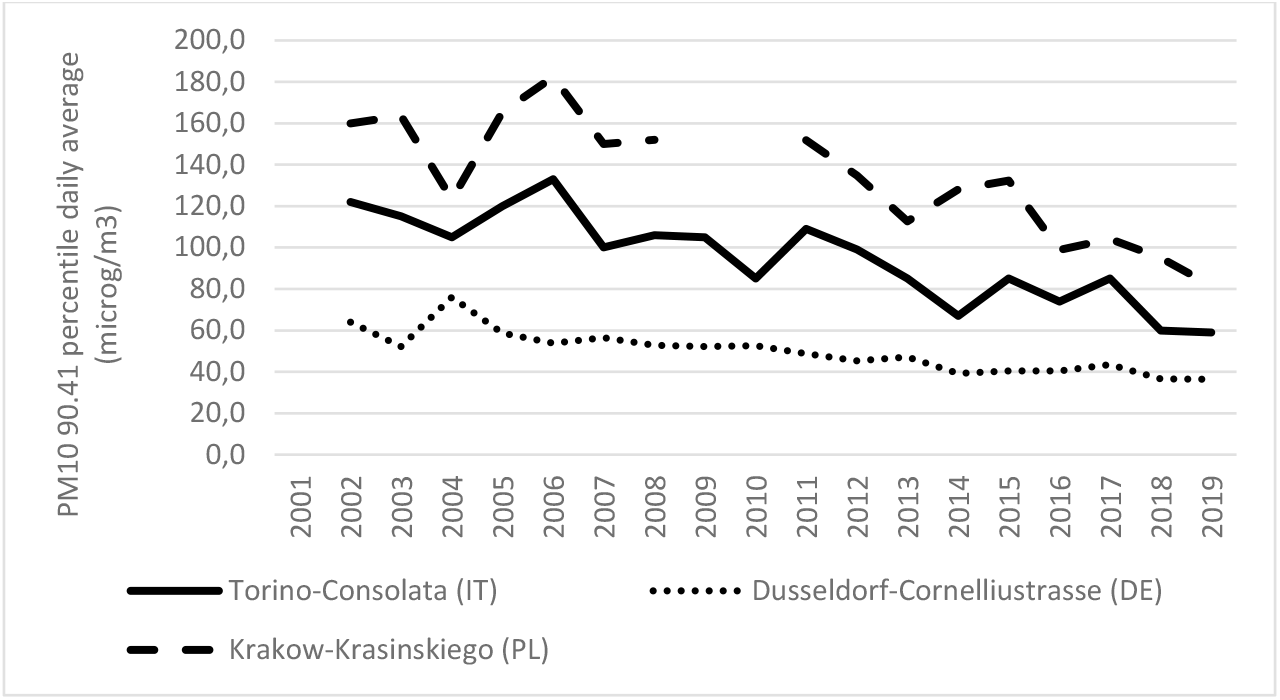
Air quality trends for PM10 (90.41 percentile daily average) measured at 3 urban-traffic stations in Torino (IT), Düsseldorf (DE) and Krakow (PL)

**Figure 6:**
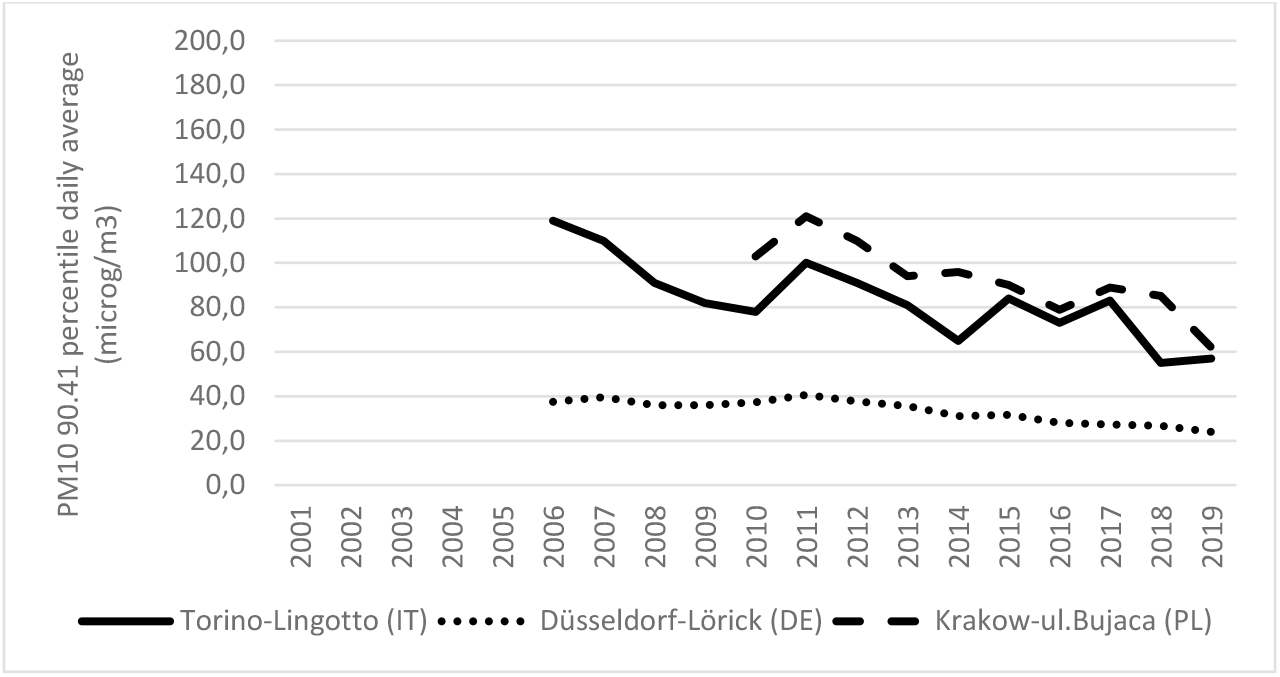
Air quality trends for PM10 (90.41 percentile daily average) measured at 3 urban-background stations in Torino (IT), Düsseldorf (DE) and Krakow (PL)

To understand the dynamics of pollutants dispersion in the atmosphere and the related chemical transformations, thus explaining the emissions-air quality relationship described in Figures 1 to 4, it is essential to analyse the meteorological parameters introduced in Section 2.2. Figure 7 shows the diurnal pattern of PBL height during the cold season in Torino (Piedmont, IT), Essen (NRW, DE) and Katowice (Silesian, PL). As the Figure points out, the PBL height in Torino as well as in the whole Po Valley is close to the ground, especially during the evening, the night and in the early morning. Pollutants emitted in this area are dispersed through a mixed layer depth comprised between 200 m and 400 m, only from 10 a.m. to 16 p.m., from December to February. In Katowice and Essen, the PBL heights are radically different, showing depths comprised between 400 and 800 m during the same cold months. Similar conditions can be found in other placed north of the Alps, such as Stuttgart, Germany or Prague, Czech Republic. In conclusion, the PBL height in Torino during the winter is four-five times lower than in Northern Europe (Figure 8).

**Figure 7:**
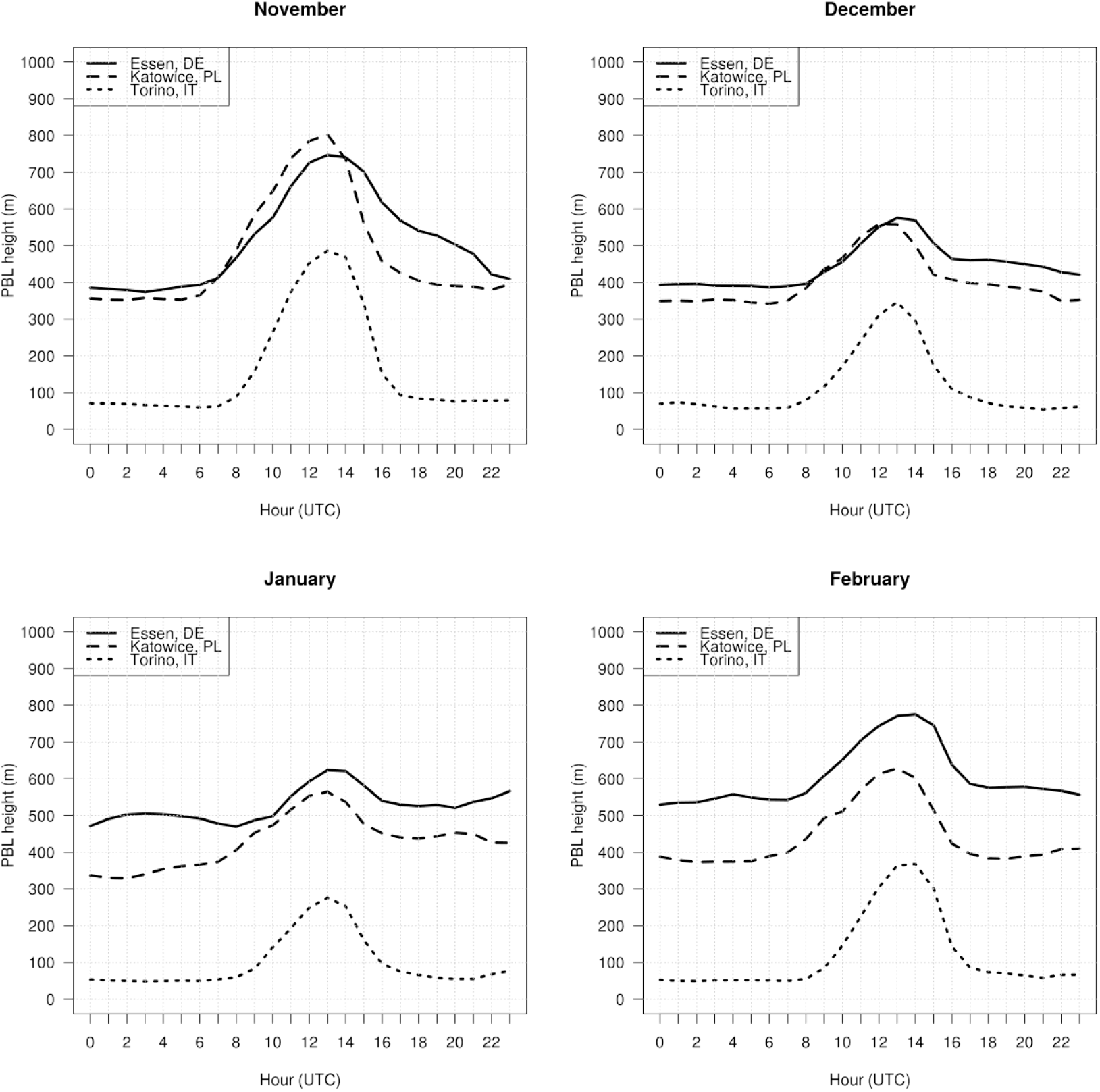
PBL height diurnal pattern in Torino (North Western Italy), Essen (North Western Germany) and Katowice (Southern Poland) during the cold season (average values from 1979 to 2020)

**Figure 8:**
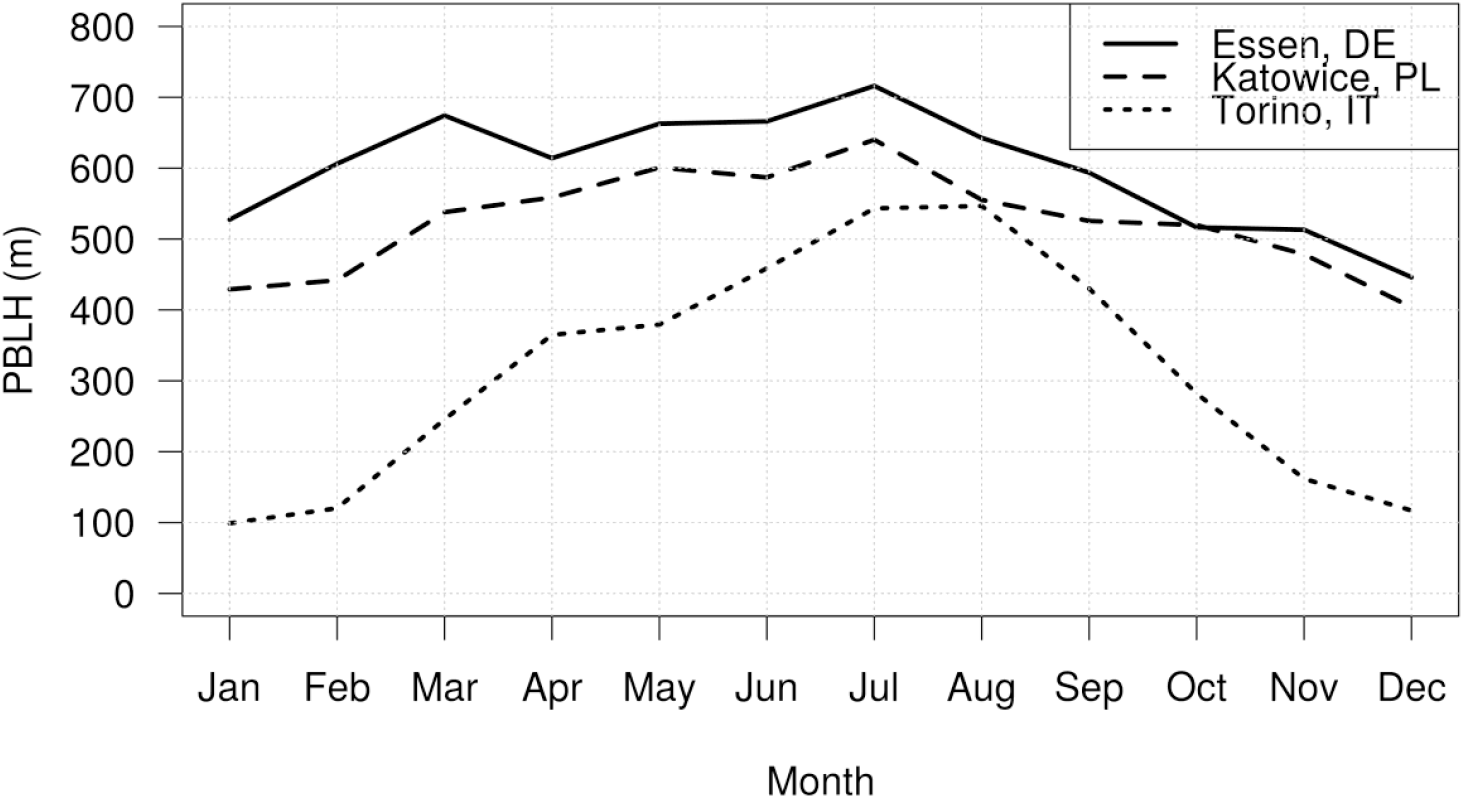
PBL height (monthly average) in Torino (IT), Essen (DE) and Katowice (PL)

As already described, PBL height is inversely proportional to PM10 concentration, as reported by Figure 9 where PM10 concentrations higher than 50 μg/m^3^ occur in Torino when PBL depth is lower than 200 m. The logarithmic fit between PBL height and PM10 concentration measured from 2011 to 2020 at the station of Torino-Consolata is statistically meaningful with slope equal to -23.3 microgr/m4 (error 1.4) and intercept equal to 170.6 microgr/m3 (error 6.5).

**Figure 9:**
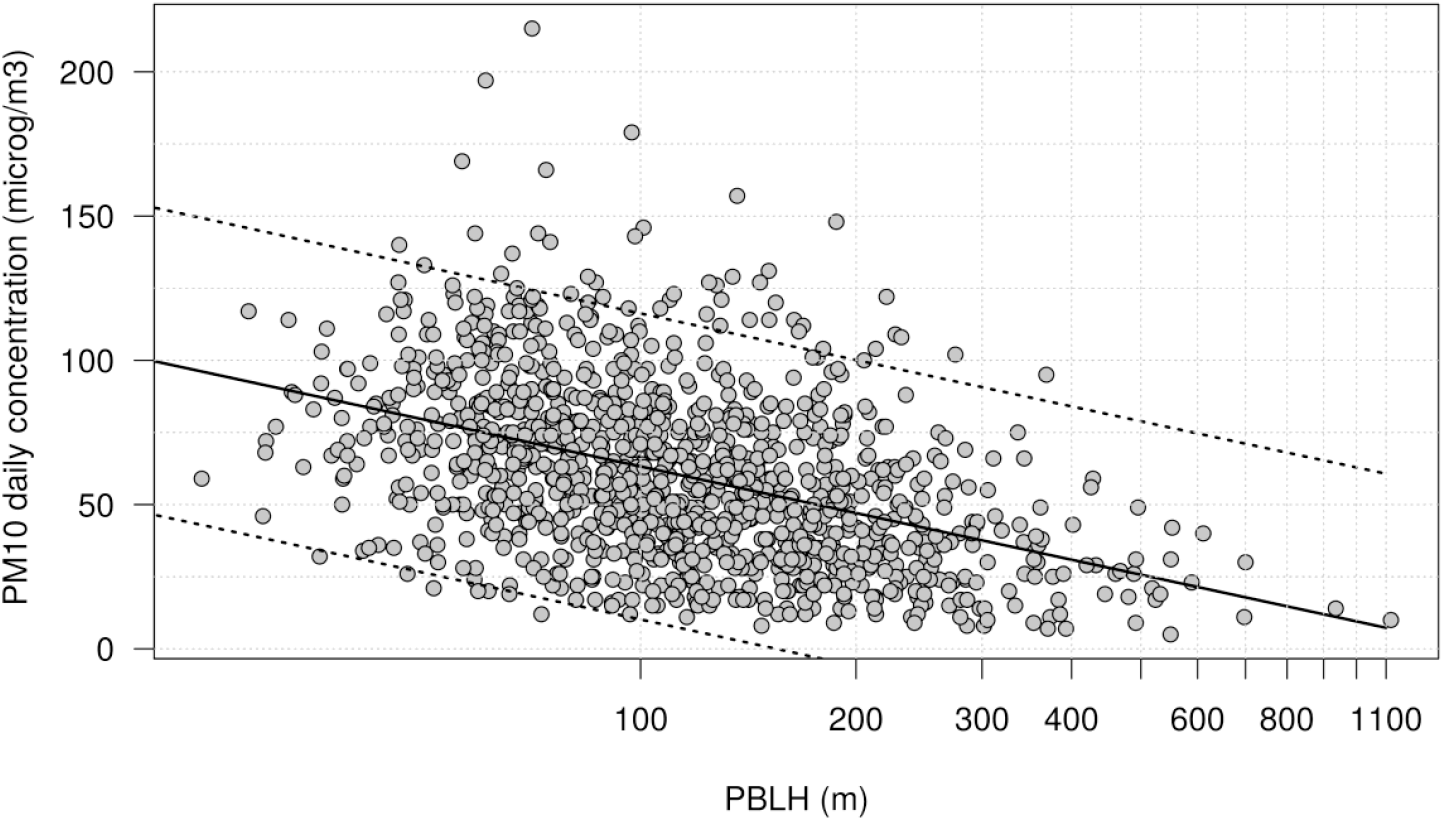
Correlation between PBL height and PM10 daily averages measured in Torino (N-W Italy) in the period 2011-2020

Contextually, Torino and the Po Valley show high-pressure values from November to February whereas in Northern Europe low pressure conditions are frequent also during the cold season, as pointed out by Figure 10. In December and January Torino shows mean sea level pressure significantly higher than those found in the other two locations. Atmospheric pressure values are the lowest in Essen during November and February, sign of dynamical atmospheric conditions, more favourable to pollutant dispersion.

**Figure 10:**
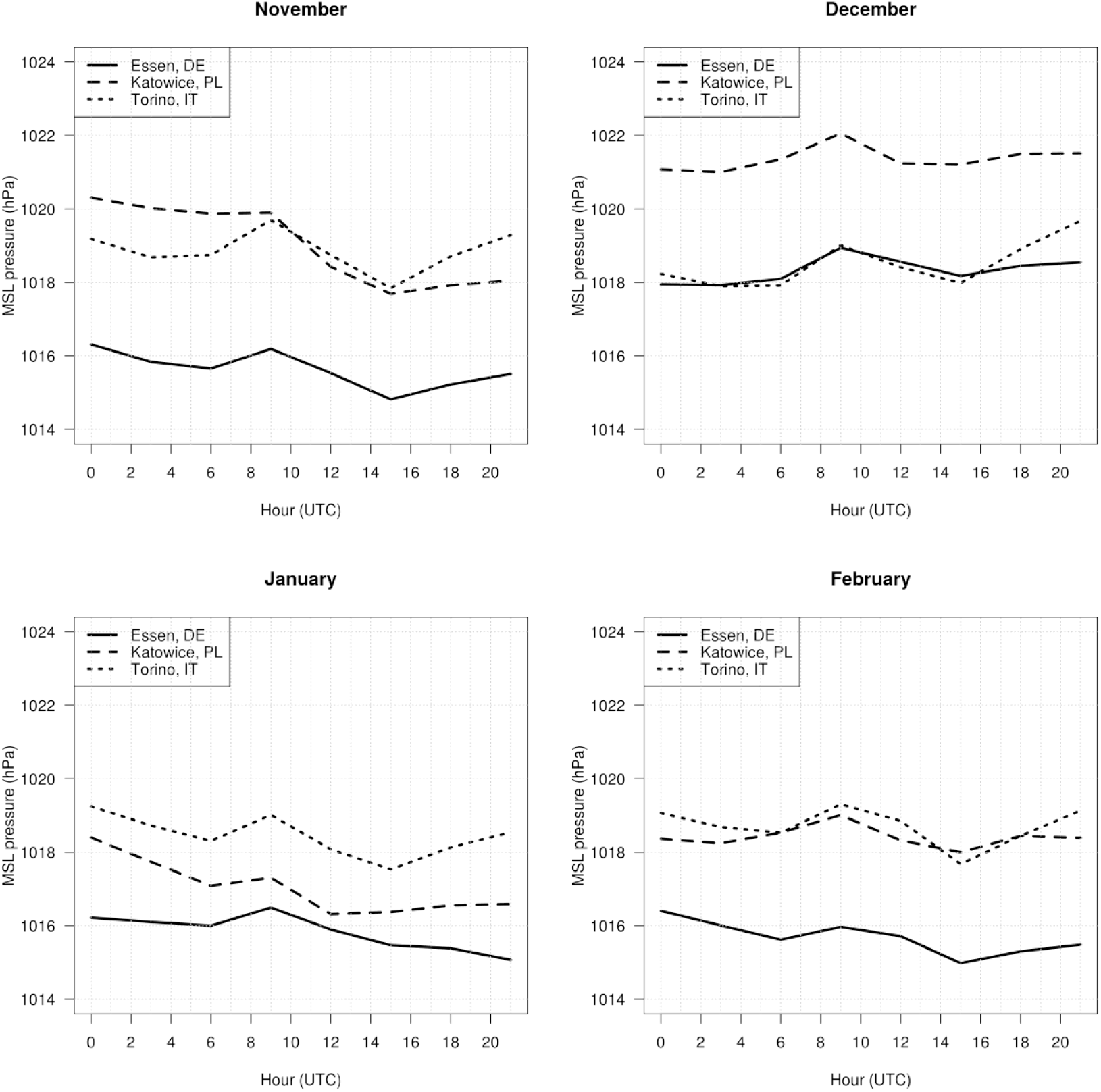
Mean sea level pressure diurnal pattern in Torino (North Western Italy), Essen (North Western Germany) and Katowice (Southern Poland) during the cold season (average values from 1979 to 2020)

Finally, Figure 11 reports the comparison of 10m-wind speed derived from ERA5 reanalysis in the three areas. Wind speed at 10 meters in Torino is always between 0.5 and 1 m/s while in Essen or Katowice wind speed is 3 to 5 times higher on average. As expected, wind speed tends to increase during the afternoon as a consequence of thermal local circulation. On February, Torino shows a slight increase in average wind speed and a less pronounced diurnal cycle; this behaviour can be interpreted as a decrease of stagnant atmospheric conditions.

**Figure 11:**
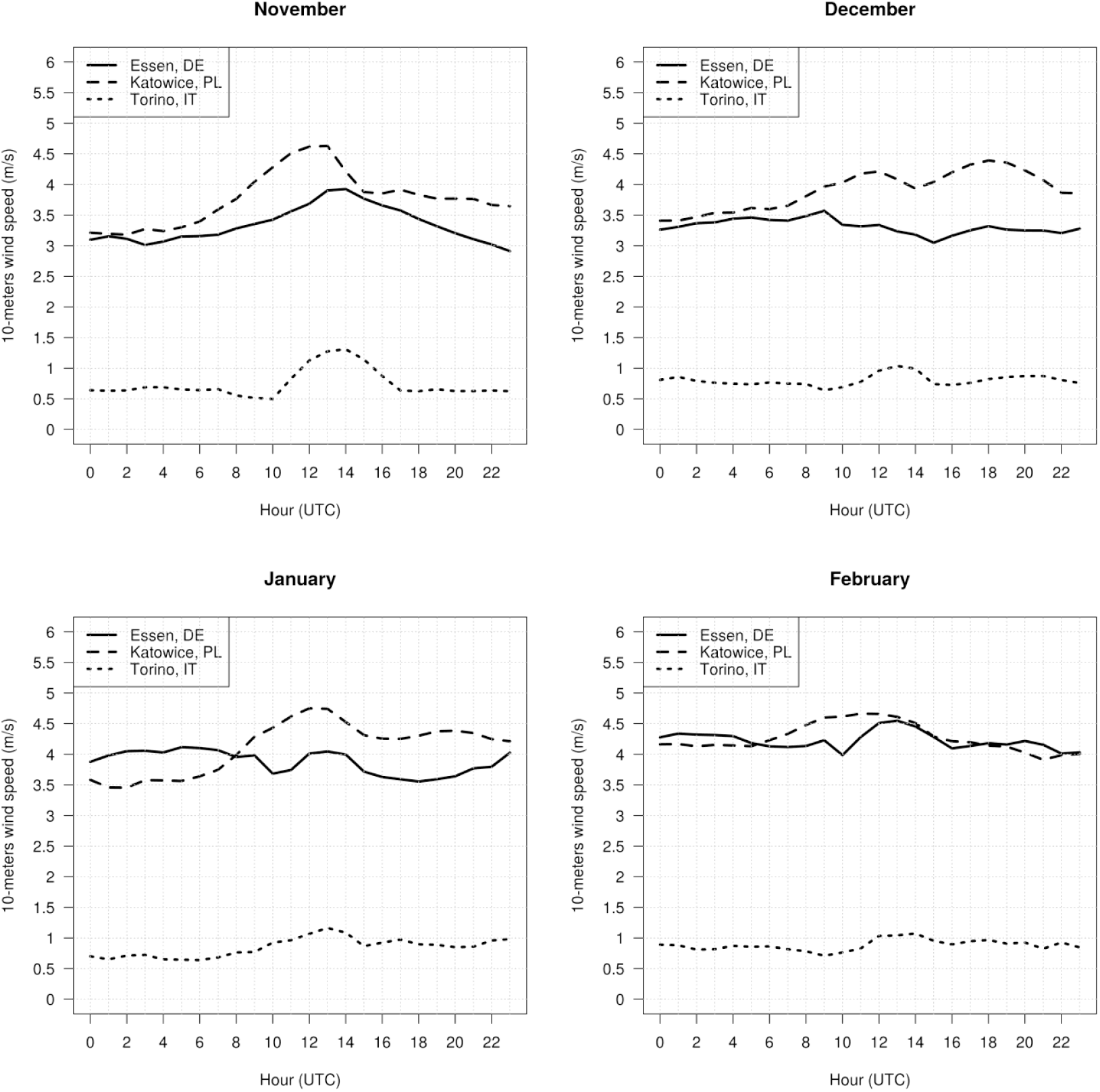
wind speed diurnal pattern in Torino (North Western Italy), Essen (North Western Germany) and Katowice (Southern Poland) during the cold season (average values from 1979 to 2020)

In conclusion, the three primary meteorological parameters, PBL height, wind speed and atmospheric mean sea level pressure, unquestionably illustrate why air quality of Northern Italy is still worse than in other places in Europe despite of the massive emissive reduction carried out in the last decades; during the cold seasons, the atmospheric dilution/dispersion capabilities are three to five times weaker than in other EU countries, favouring stagnation of pollutants and chemical transformation of PM10 gaseous precursors.

This last aspect can be developed and discussed through PM analytical Source Apportionment described in Chapter 2.3; this method can determine PM main constituents and, consequently, main emissive contributors. Figure 12 reports mass contributions to PM10 concentration as calculated for Milano-Pascal and Torino-Lingotto air quality stations in Northern Italy: as pointed out by the plots, in the Po basin, secondary inorganic aerosol (ammonium sulphates and nitrates) represents, on average during the cold season, the largest contribution to PM10 concentration. SIA can represent more than 50% of PM10 concentrations during the critical episodes of air pollution (Lanzani, 2017). The contribution of SIA is homogeneous over the Po Valley (Scotto et al., 2021).

**Figure 12:**
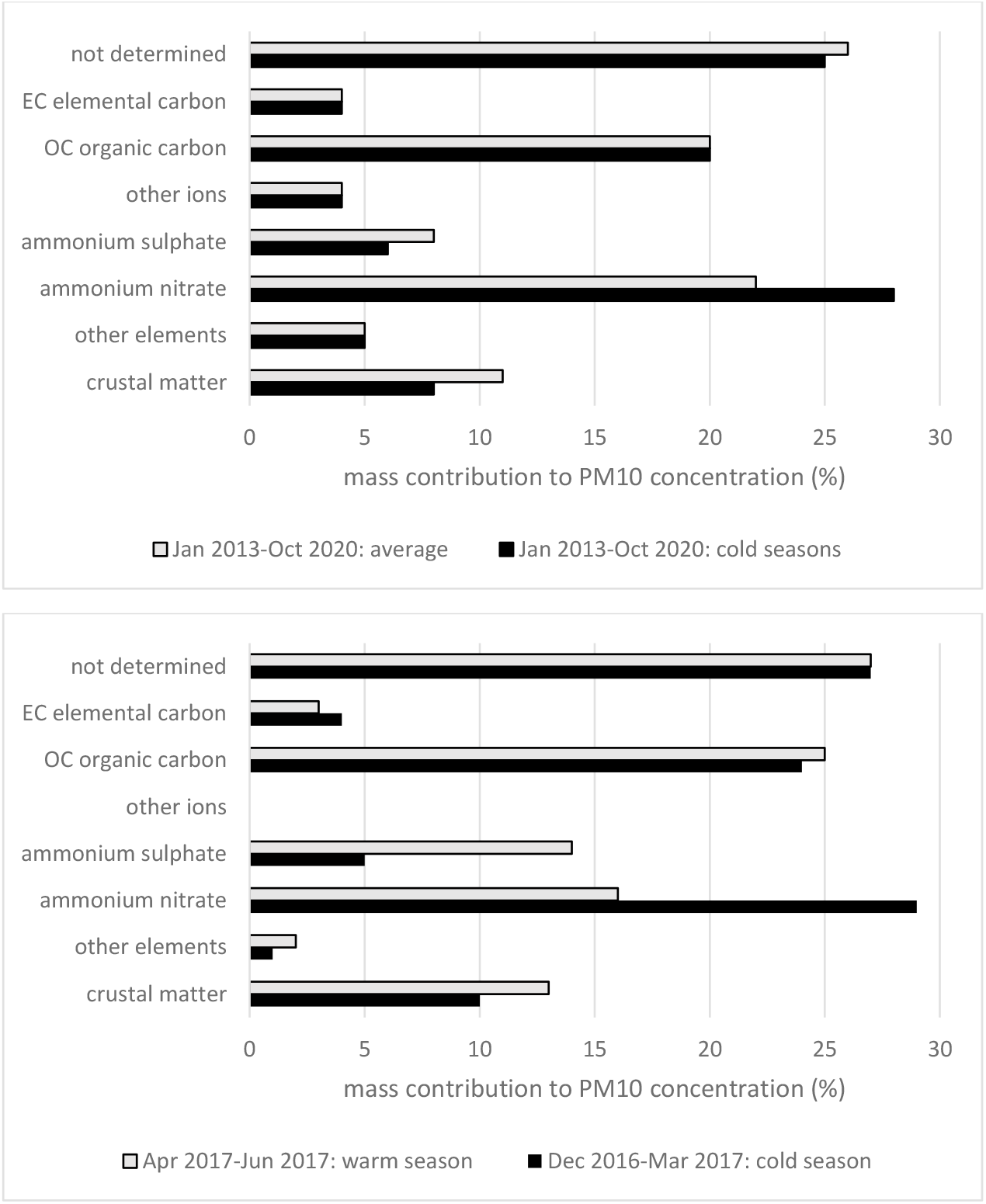
PM10 constituents form analytical Source Apportionment in the Po Valley (for Milano-Pascal station in the upper plot and for Torino-Lingotto in the lower one)

Provided that secondary inorganic fraction of PM is of primary importance in the Po basin, it’s worth remembering the main patterns followed by atmospheric chemical transformation of gaseous pollutants to PM. Ammonia is *de facto* the only base in the gas phase in our atmosphere. It rapidly reacts with the available acids (mainly sulfuric and nitric, coming from SOx and NOx emissions) to form the corresponding salts. Ammonia reacts first with sulfuric acid, to form bisulphate, and, if enough ammonia is present, ammonium bisulphate. If ammonia concentration exceeds the stoichiometric threshold (twice the sulphate in moles) then some free ammonia is available to react with other acid gases too, such as nitric acid, to form ammonium nitrate (Seinfeld and Pandis, 2006). While ammonium sulphate is a relatively stable compound, ammonium nitrate is not. Ammonium nitrate tends to evaporate (the reaction is reversible) and its formation is favoured by low temperatures and high relative humidity, typical conditions for the Po Valley.

While in the Po basin, after a national desulphurization campaign, SOx is emitted in small quantities with respect to ammonia and NOx, that is there is free ammonia reacting with nitric acid to form ammonium nitrate, in Northern Europe, where sulphur containing fuel is massively used and SOx emissions are large (see for example North Rhine-Westphalia or Southern Poland emission inventory), ammonia neutralizes first sulphuric acid and NO2 is less “consumed” to form secondary particles, thus resulting in higher concentrations (as reported by Figure 1).

Indeed, PM Source Apportionment carried out in Northern or Central Europe shows very different constituents’ profiles with respect to Northern Italy. Weijers et al. (2011) reports that in an urban-background station placed in Schiedam (part of Rotterdam (NL) urban agglomeration with appr. 600,000 inhabitants), PM2.5 is made up of ammonium sulphate for 24%, ammonium nitrate for 22%, Elemental Carbon for 14%, Organic Carbon for 10%.

Shen et al. (2019) determined the composition and origin of PM2.5 aerosol particles in the upper Rhine Valley (South-Western Germany) during the summer, showing that the main contributions is referable to organic matter, followed by Sodium salts; contribution of secondary inorganic aerosol is not decisive.

Another German research group (Hellack et al., 2015) analysed 75 PM10 samples collected at an urban background station in Mülheim-Styrum, North Rhine-Westphalia. Here, 7 contribution factors were identified, namely mineral dust (that is crustal matter), secondary nitrate and sulphate, industry, fossil fuel combustion, non-exhaust traffic and marine aerosol. For the marine aerosol factor, higher contributions are due to western and northern derived air masses, in contrast, fossil fuel combustion factor corresponds to eastern winds; long distance PM transport might lead to enlarged contribution of the mineral dust and fossil fuel combustion from continental, eastern European areas. The eastern driven fossil fuel combustion factor could be additionally due to emissions from the industrial Ruhr area. During the fall and winter, the main contribution to PM composition is attributed to industry and fossil fuel combustion.

As far as Poland is concerned, a first study focused on PM2.5 collected in Warsaw for a full year (Juda-Rezler, 2020) reports that Organic Carbon contributes for 29.6% to particulate matter annual concentration, Elemental Carbon for 7.8%, SIA is 30.7% (70% of SIA is made of ammonium sulphate), elements 4.4%, other ions 5.2% and the remaining 22.3% is unidentified.

Another Polish study (Samek et al., 2017) describes a Source Apportionment campaign carried out in Krakow, where combustion and biomass burning account respectively for 22.9 and 15.6% of the annual PM2.5 concentration, sulphates and nitrates represent 19.3 and 17.1% and traffic 8.3%.

A European-wide overview of PM Source Apportionment study is available from Viana et al. (2008), although data are not recent. Here, different contributions of crustal elements, sulphates from industrial activity, carbonaceous matter and SIA can be appreciated for many European sites, from Portugal to Finland. Lastly, Almeida et al. (2020) produced a very interesting Source Apportionment study involving 16 European and Central Asia urban areas.

Provided that PM composition of Northern Italy could be quite different if compared to other European regions, as pointed out in the previous paragraphs, also the effect of measures aiming at reducing emissions could be different from what is expected. In this sense, emission reductions due to Covid-19 lockdown can be used as a real experiment in order to observe the effectiveness of possible policies on air quality.

Concerning primary PM10, the total quantities emitted in Piedmont (N-W Italy) up to the first half of April 2020 remained substantially unchanged with respect to those occurring without lockdown, as the reduction of the contribution from industry and traffic was offset by the overall increase in domestic heating emissions. On the other hand, in the case of nitrogen oxides, a net reduction in emissions, up to 30%, is observed compared to the same period of an ordinary year, as the prevalent contribution to NO2 emissions is given by vehicular traffic.

In Figure 13, NO2 concentrations measured at the urban-traffic station of Torino-Consolata (N-W Italy) shows a sharp decline in the month of March and April 2020 compared to the first two months of the average year.

**Figure 13:**
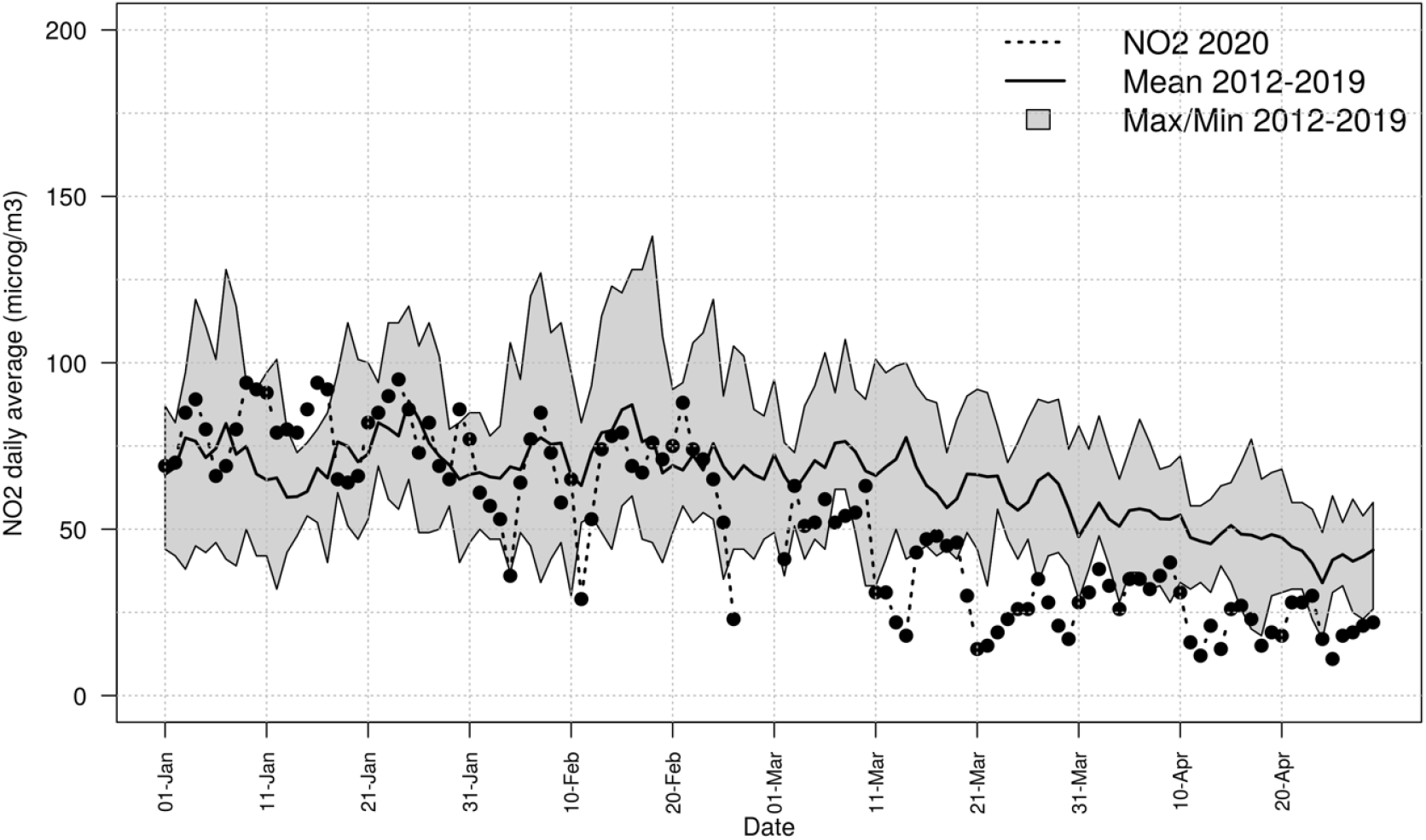
NO2 concentrations measured at Torino-Consolata station (urban-traffic) during Covid-19 lockdown of March and April 2020, compared to historical data (2012-2019)

Figure 14 reports PM10 concentrations measured at the urban-background station of Torino-Lingotto (N-W Italy); the plot highlights that, even though emissions of PM precursors (NO2 in particular) have been strongly diminished by lockdown, in March and April 2020 the daily PM10 concentrations lay generally within the variability range of the reference period (with some episodes of very high concentrations). At the end, 2020 in Piedmont turned out to be worse than 2019 for PM10 daily concentration exceedances because of fewer rainfall episodes.

**Figure 14:**
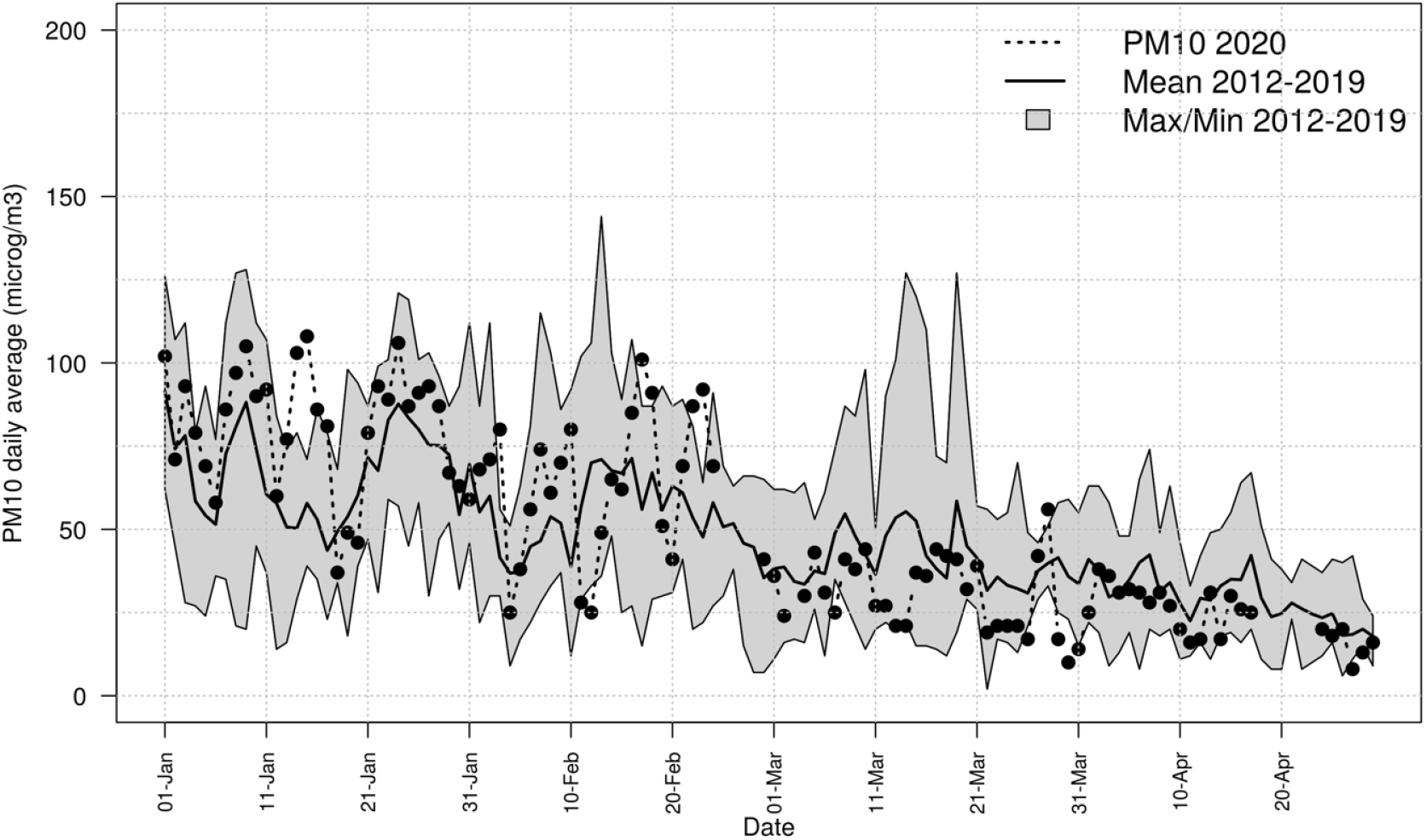
PM10 concentrations measured at Torino-Lingotto station (urban-background) during Covid-19 lockdown of March and April 2020, compared to historical data (2012-2019)

In the same way, the third report by PREPAIR project (EU LIFE IP Prepair, 2021) investigates the lockdown effects on air quality in the entire Po Valley. Here maximum reductions of NOx and primary PM10 emissions of 40% and 20% have been assessed; in the face of a general reduction of gaseous pollutant concentration observed during the lockdown, PM concentrations didn’t decrease, showing positive and negative fluctuations mostly depending on meteorological conditions. The report observes a strong reduction of Elemental Carbon contribution to PM during the lockdown due to traffic reduction, as well as a reduction of elements deriving from industrial activities. At the same time, contributions due to biomass burning for residential heating increase and, most of all, secondary inorganic aerosol were even higher or invariant with respect to 2019 and historical series. The hypothesis that can explain the behaviour observed for PM10, according to the cited study, is that despite of a strong reduction of NOx emissions, the main precursors of SIA, in particular ammonia, were present in sufficient quantities to support secondary aerosol formation during lockdown. In other words, NOx concentrations do not seem to behave as a limiting factor for ammonium nitrate formation in the Po basin.

## 4. Discussion

### 4.1 Difficulties to comply with air quality limits in Northern Italy

The previous chapters described the results of analyses aiming at understanding air quality conditions of the Po Valley, starting from i) the drivers, that are areal emissions, ii) the meteorological parameters influencing atmospheric pollutants’ dispersion and transport, iii) the composition of PM and iv) the effectiveness of strong emissive reductions on air quality during lockdown periods.

The discussed data show that concentrations of atmospheric pollutants in the Po Valley had one the best reduction trend in Europe in the last 20 years, due to very large emissive reduction, again among the highest in Europe, as pointed out by figures from EEA. Now, in the face of lower or at most comparable emissions, the Po basin regions still have PM10 and PM2.5 noticeably worse than other European regions placed, for example, in Germany.

The reasons why the Po basin still exceeds EU air quality limits (as well as, obviously, the stricter WHO guidelines levels) are normally attributed to orography and meteorological adverse conditions. The aim of the elaborations described at chapter 2.2 is to prove, objectively, that atmospheric dispersion and dilution capacity of Po basin is drastically lower than elsewhere in Europe as planetary boundary layer height is often lower than 200 m during the cold seasons, 5 times less than in Northern Europe.

When pollutants remain concentrated in a smaller volume and wind is too weak to transport pollutants elsewhere, contrary to what usually happens in Northern Europe where transboundary pollution is an important issue, stagnation favours chemical transformation of gaseous pollutant toward particulate and higher precursor concentrations lead to higher reaction kinetics for secondary pollution. During accumulation episodes, when primary pollutants tend to accumulate near the ground, reaching high concentrations and therefore favouring the formation of further secondary particles, pollution is no longer limited to urban areas, but high concentrations are usually measured all over the Po plane, also in rural areas.

The described conditions are confirmed by Source Apportionment campaigns developed in Northern Italy, where secondary inorganic aerosol (SIA) is the main contributor to PM composition: indeed, SIA could account for more than 30% of PM10 on an annual average and, during the peak episodes in the winter it can represent up to 70% of PM10. PM components’ profile for the Po Valley could be very different compared to other regions in Europe, in particular north of Alps; contributions of Elemental Carbon (EC) and ammonium sulphate is in fact lower in the Po basin, while a high concentration of ammonium nitrate can be found, in particular during the cold seasons.

As a consequence of what we briefly summarized in these paragraphs, not even radical emissive reductions such as those occurring during Covid-19 lockdown in the late winter/spring of 2020 can reduce PM10 concentrations as one could expect as the extinction or the reduction of a part of the pollutants is not sufficient to cause an appreciable change in secondary aerosol formation.

Another very interesting study (Raffaelli et al., 2020) confirms these results. Here, the research group tries to assess the emissive reductions to be reached in the Po basin to comply with EU air quality limits for PM10 (daily concentration of 50 μg/m^3^, to not exceed for more than 35 days a year) and NO2 annual mean value (40 μg/m^3^). The evaluation is carried out by using a complex modelling suite managed by the Environmental Agency of the Emilia-Romagna Region (ARPAE). The research establishes an “action-plans scenario” where the emission reductions are very important for all pollutants: NOx decrease should be 39%, PM10 38%, PM25 40%, NH3 22% and SO2 3%, as compared to 2013 emissions. The emissive reductions rely mainly on improvements to be carried out by mobility for NOx, domestic biomass burning for PM and agriculture for NH3.

The calculated necessary reductions correspond to 30,000 t/y of primary PM10, 150,000 t/y of NOx, 54,000 t/y of NH3, that represents a big challenge, maybe unrealistic with the present technologies, territory, population, fuels, and habits. 150,000 t of NOx correspond in fact to the whole NOx emissions of 2 Northern Italy regions, Piedmont and Emilia Romagna, accounting for more than 8.7 million inhabitants, or Bavaria in Germany; 30,000 t of PM10 account for the emissions from 9 million people living in Piedmont and Veneto, more than Austrian PM10 emission, that are around 26,000 t/y; emissions of 54,000 t/y of ammonia means the elimination of current agricultural and farming emissions from Veneto or Baden-Württemberg.

The same research group affirms that an emissive reduction at this level would have considerable economic and social impacts, introducing an “element of disparity” towards Northern Italy competitivity and society. Further restrictions of air quality limits as those under study by European Commission would require even larger emissive reductions.

On the other hand, as reported by Raffaelli et al., the same baseline emissive scenario modelled for the month of December 2018 in central Europe, with much more favourable atmospheric dispersion conditions, could lead to lower PM10 and NO2 concentrations from 60 to 80% compared to Northern Italy. Interestingly, this result, obtained by a complex modelling approach, is the same order as the outcome of a simple box model, where the same emissions are dispersed in a volume three to five times smaller because of a substantially lower PBLH, as discussed in chapter 2.2.

This way, as far as the compliance with air quality limits in Northern Italy is concerned, a possible research target could be an overall cost-benefit analysis, where costs are associated to the application of measures from air quality plans and benefits can be defined as avoided external costs involving air pollutants. CAFE-CBA methodology (http://europa.eu.int/comm/environment/air/cafe/activities/cba.htm) has been developed and applied to quantify damages to health and crops due to units of pollutant release (AEA Technology Environment, 2005). The results of the cited study show very large variations between countries concerning damages per tonne emission. Generally, the highest damages are found for emissions occurring in central Europe and the lowest for countries around the edges of Europe. This simply reflects variation in exposure of people and crops to the pollutants of interest that is at most at the centre of Europe.

Indeed, provided that very differentiated situations concerning emissions, air pollution dispersive conditions and air quality occur all over Europe, regional, national and European Union funds could be managed and addressed according to the actual atmospheric dispersion potential of European regions, so as to rebalance the evident disadvantages depending on orography and meteorology and to finance air quality measures where they are at most necessary.

It is worth pointing out that substantial improvements of air quality could be reached through strong measures involving biomass burning (to reduce primary PM10) and agriculture (to reduce ammonia emissions that could act as a limiting factor to inorganic secondary PM formation). In this regards, penetration and respect of air quality measures imposed by regional or interregional plans is an important issue. Firstly, the application of best available techniques for farming and manure spreading is far to be reached, at least as far as we know. Secondly, we must say that if biomass stoves are not carefully maintained, measures enforcing the use of high-quality pellets (according to European standard EN ISO 17225-2) in high performance heating plants (4 or 5-star according to the classification introduced by Italian Environmental Minister Decree D.M. 186/2017) would be useless to improve air quality as emissions from stoves would be the same as those produced by old stoves fed by low-grade biomass fuel: Table 3 reports the results of a real emission study carried out by Arpa Piemonte on a 4-star stove fed by pellet classified as A1 (the highest quality according to EN ISO 17225-2). A possible solution to air quality problems arising from biomass could be represented by the use of biomass as a fuel for district heating systems, where emissions are adequately reduced by abatement systems and controlled, instead of a widespread biomass combustion in very small stoves or domestic burners.

**Table 3:** Emissions from a 10.8 kW stove fed by high quality pellet (A1 according to EN ISO 17225-2). Concentrations are expressed on dry gas, O_2_ at 13%; EN sampling/analytical reference methods have been applied

|  | CO<br>mg/Nm<sup>3</sup> | NO<sub>x</sub><br>mg/Nm<sup>3</sup> | VOC<br>mgC/Nm<sup>3</sup> | PM<br>mg/Nm<sup>3</sup> | CH<sub>2</sub>O<br>mg/Nm<sup>3</sup> | PAH<br>µg/Nm<sup>3</sup> |
| --- | --- | --- | --- | --- | --- | --- |
| pellet stove with regular maintenance | 331 | 98 | 4,3 | 20 | 0,5 | 0,39 |
| pellet stove without regular maintenance | 537 | 97 | 21,7 | 63 | 0,8 | 4,3 |
| 4-star stove certified emissive performance* | 250 | 160 | 35 | 20 |  |  |
| 2-star stove certified emissive performance* | 500 | 200 | 80 | 50 |  |  |

\* according to the Italian Environmental Minister Decree D.M. 186/2017

### 4.2 PM toxicity and air quality standard

Since the health burden due to PM-related air pollution is one of the biggest environmental health concerns, limit values and policies should be imposed based on a thorough knowledge of health effects. In 2007, a WHO report (WHO Europe, 2007) stated that: <<In the future, better understanding of the relative toxicity and health effects of particles from various sources could facilitate targeted abatement policies and more effective control measures to reduce the burden of disease due to air pollution.>>. At that time, monitoring data on component-specific PM concentrations were scarce as well as relevant exposure data, moreover existing inventories suffered from gaps in emission data; on the contrary, consistent evidence for the association of PM emitted by the major combustion sources, mobile and stationary, with a range of serious health effects, including increased morbidity and mortality from cardiovascular and respiratory conditions, was already at disposal. Different chemical characteristics of particles, the report declared, have different relative risks on a per-unit-mass basis; in particular, one of the reported hypotheses was that the oxidative potential of the particles or specific components (for example, transition metals and combustion-derived primary and secondary organic particles) could be one of the PM’s mechanisms of action because of the greater ability to deplete antioxidant defences. The knowledge at disposal, however, didn’t allow in 2007 precise quantification or definitive ranking of the health effects of PM emissions from different sources or individual PM components.

In 2013 a new WHO report (WHO Europe, 2013) confirmed that: a) new evidence links black carbon particles (that is Elemental Carbon) with cardiovascular health effects and premature mortality, for both short-term (24 hours) and long-term (annual) exposures; b) concerning secondary inorganic aerosol, neither the role of the cations (for example, ammonium), nor the interactions with metals or absorbed components have been well documented in epidemiological studies; c) there is growing information on the associations of organic carbon with health effects.

Other following studies (Cooke et al., 2007; Tuomisto et al., 2008; Lelieveld, 2015) clearly focused on health impact of outdoor air pollution depending on assumptions on toxicity of particles.

In 2021, the last report by WHO (WHO, 2021) affirms textually that: << many studies have tried to identify which sources and/or physicochemical characteristics of airborne PM contribute most greatly to toxicity. This is a challenging area of research, given the great heterogeneity of airborne particles, and a definitive set of particle characteristics has yet to be identified>>.

At the same time, Lelievel et al. (2020) applied an interesting modelling tool (Global Exposure Mortality Model) derived from many cohort studies to assess excess mortality attributable to ambient air pollution (PM2.5 and ozone) on a global scale. Here, the loss of life expectancy (LLE) due to air pollution at a European scale is evaluated 2.21 years, where LLE for France is 1.63, 1.91 for Italy, 2.41 for Germany, 2.77 for Czechia and 2.83 years for Poland among the other countries. The reported study points out that the fraction of avoidable LLE attributed to fossil fuel is nearly two-thirds globally, and up to about 80% in high-income countries.

Another recent scientific report (Park et al., 2018) focuses on toxicity of fine particles produced from various combustion sources (diesel engine, gasoline engine, biomass and coal combustion) and non-combustion sources (road dust, sea spray aerosols, ammonium sulphate, ammonium nitrate, and secondary organic aerosols (SOA)), so as to obtain toxicity scores for different PM components through source-specific toxicity test. The study determined multiple biological and chemical endpoints (oxidative potential (OP), cell viability, genotoxicity (based on mutagenicity and DNA damage), oxidative stress and inflammatory response). Interestingly, higher toxicity was assessed for combustion aerosols, in particular diesel engine exhausts, compared to non-combustion PM. In particular, genotoxicity (mutagenicity) and OP of diesel engine exhaust particles (i.e. soot) were significantly higher than those of other aerosol types probably because of the presence of organic components (e.g., PAH) able to break DNA strands through reactive oxygen species (the so-called ROS). Biomass burning showed toxicity comparable to that of diesel engine exhaust particles. Toxicity decreases for bituminous coal combustion (carried out at high temperature), resuspended road dust, desert dust. Ammonium sulphate/nitrate showed little toxicity whereas SOA toxicity turned out to be comparable to that of biomass burning. In order to obtain a toxicity ranking for different source PM, the authors applied the Correlation Coefficient and Standard Deviation (CCSD) method which attributed the highest differential weight to the endpoint cell viability, followed by mutagenicity, oxidative potential, inflammatory response, and oxidative stress.

The study obtained the normalized toxicity scores (0 to 10) for source-specific aerosols reported in Figure 15.

Since the authors declare that source-specific toxicity scores are additive, by combining the toxicity scores for source-specific aerosols with the mass fractions of sources in ambient particulates determined by PM source apportionment, a toxicity factor for ambient PM10/PM2.5 could be obtained as follows:

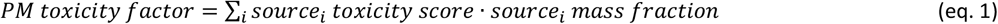

Since mass fraction are expressed as percentage and toxicity score ranges between 0 and 10, PM toxicity factor calculated according to equation 1 falls within the interval 0-10.

**Figure 15:**
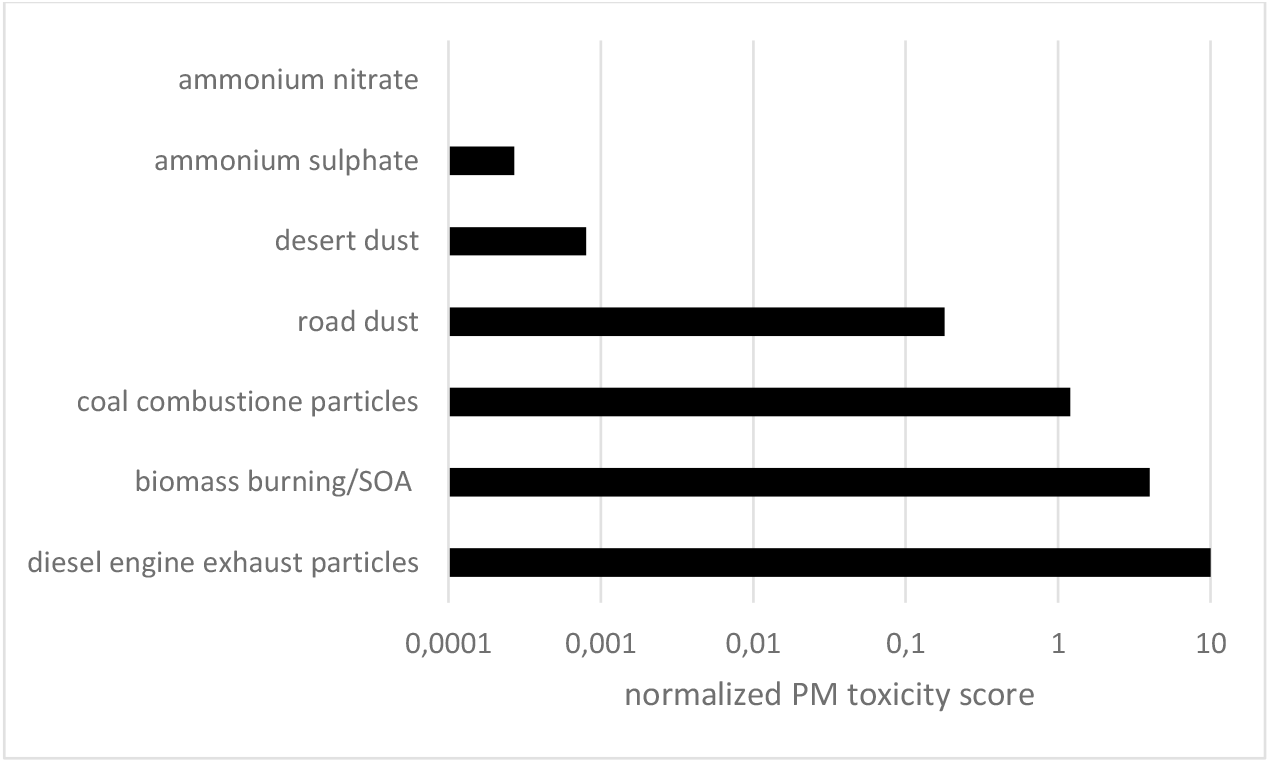
Normalized toxicity score (0 to 10) for source-specific aerosol with differential weights given to the endpoint (data taken from Park et al., 2018)

Thus, the toxicity scores proposed by Park et al. (2018) have been coupled with the Source Apportionment results illustrated in chapters 2.3 and 3 (EU LIFE IP Prepair, 2020; Weijers, 2011; Shen, 2019; Hellack, 2015; Juda-Rezler, 2020; Samek, 2017; Viana, 2008; Almeida, 2020), involving many European and Central Asia urban areas. The obtained PM toxicity factors ranges from 0.3 (for areas where the main PM contribution is referable to sea salts or mineral matter) to 3.5 (where Elemental and Organic Carbon prevail), suggesting that, even at the same mass concentration, the effects of PM10/2.5 on human health are significantly variable and limit values should take into account differential toxicity.

Park et al. conclude by saying that the <<knowledge of toxicity of particles produced from various sources obtained here can be linked with Source Apportionment and exposure level to derive a new health metric for ambient PM2.5 in future work. Differential toxicities of particles provide information that is more relevant for decision makers to establish PM2.5 abatement policies rather than only focusing on PM2.5 mass concentration>>. Provided that PM components, and linked public health impacts, are strongly variable across Europe, we totally agree with the statement of this scientific report as well as the recommendations by WHO. As a matter of fact, if a far higher toxicity of Elemental Carbon and Organic Carbon coming from combustion (traffic, biomass and solid fuel combustion) compared to secondary inorganic aerosol or dust resuspension could be confirmed, policies would be focalized, for example, on biomass burning or diesel vehicle emissions, to consistently maximise the effectiveness of health protection measures.

After all, many environmental limit values or guidelines are based on the concept of “equivalent toxicity”: the “Toxic Equivalent” (TEQ) scheme weighs the toxicity of the less toxic compounds as fractions of the toxicity of the most toxic one. For example, limits for PCDDs, PCDFs and PCBs work on that scheme. Moreover, according to Directive 2008/50/EU, air quality limits are already imposed on the most toxic heavy metals (As, Pb, Ni, Cd), PAH (benzo(a)pyrene) and VOC (benzene), confirming that regulation should take toxicity into account.

Therefore, new health metrics for ambient PM could overcome the shortcomings of the current regulation standard helping regional authorities to properly manage air quality questions, minimizing related health impact.

## 5. Conclusions

The present paper reports an in-depth analysis of the reasons why the regions of the Po valley, Northern Italy, still have difficulties to comply with EU air quality standards, in particular for PM10 and NO2, in spite of strong emission reduction carried out through careful Air Quality Plans put in practice during the last 2 decades.

The analysis includes a consistent comparison of emission inventories for different European regions in Italy, Germany and Poland, the measured air quality trends in these areas and, most of all, a thorough investigation of meteorological parameters influencing atmospheric pollutant dispersion and transport. The study reports that in the colder seasons, wind speed, PBL height and atmospheric pressure occurring in the Po basin are three to five times less efficient in diluting and dispersing pollutant if compared to regions north of the Alps. As a consequence, also against lower emissions and stronger emissive reduction trends compared to other European regions, the Po basin still has PM10 and PM2.5 noticeably worse than elsewhere. It has been demonstrated by EU LIFE-IP PREPAIR project that only radical emission reductions could bring air quality into EU standards (or stricter guidelines values), causing considerable economic and social impacts towards Northern Italy competitivity.

However, we must consider that air quality standards (particularly for PM10 and PM2.5) aim at protecting people from adverse health effects arising from air pollution. Even though in 2019 Northern Italy regions have among the highest life expectancy in Europe (83.3 years for Piedmont (IT), 84.2 for Lombardy (IT), 80.9 for North Rhine-Westphalia (DE), 82.3 for Baden-Württemberg (DE), 74.0 for Silesian (PL), according to Eurostat data), healthy air quality represents an issue to be properly addressed.

In this regard, it is necessary to also consider the toxicity of atmospheric particulate in addition to PM10/PM2.5 mass concentration as a limit value, as already pointed out by WHO reports and many toxicological studies. In fact, based on the Source Apportionment studies at disposal, on the annual average more than 40% of PM10 in the Po Valley is made up of secondary inorganic aerosol and crustal matter, that constitute PM components with lower toxicity compared to organic matter coming from traffic (diesel engine exhaust, in particular) and solid fuel combustion.

Modern PM Source Apportionment techniques, along with reliable toxicity and epidemiological analyses, represent the right tools to build a new consistent health metric for ambient PM in the future, helping policy makers impose effective air quality measures to protect people health.

## Data Availability

All data produced in the present study are available upon reasonable request to the authors

## Acknowledgements

The authors acknowledge the scientific support coming from EU LIFE-IP Po Regions Engaged to Policies of Air (PREPAIR) project and the precious analysis of the contributing scientists.

